# Prevalence of diabetic peripheral neuropathy in Africa: A systematic review and meta-analysis

**DOI:** 10.1101/19007401

**Authors:** Wondimeneh Shibabaw Shiferaw, Tadesse Yirga, Yeshamble Work, Yared Asmare Aynalem

## Abstract

**Introduction:** Diabetes mellitus is a global health care problem and financially costly. Diabetic peripheral neuropathy is common and frequent cause of morbidity and disability. Despite its serious complications, limited evidence is available on the magnitude of diabetic peripheral neuropathy among patient with diabetes mellitus. Hence, the objective of this systematic review and meta-analysis was to estimate the pooled prevalence of diabetic peripheral neuropathy among patients with diabetes mellitus in Africa.

**Methods:** PubMed, Scopus, Google Scholar, Africa journal online, WHO afro library and Cochrane review were systematically searched online to retrieve related articles. The Preferred Reporting Items for Systematic Review and Meta-Analysis (PRISMA) guideline was followed. Heterogeneity across the included studies was evaluated by inconsistency index (I^2^). Publication bias was examined by funnel plot and Egger’s regression test. The random-effect model was fitted to estimate the pooled prevalence of diabetic peripheral neuropathy among diabetes mellitus patients. All statistical analysis was done using STATA version 14 software for windows.

**Results:** Twenty-three studies which comprises of 269,691 participants were included in the meta-analysis. The overall pooled prevalence of diabetic peripheral neuropathy was 46% (95% CI:36.21-55.78%). Based on the subgroup analysis, the highest magnitude of diabetic peripheral neuropathy was reported in West Africa 49.4% (95% CI: 32.74, 66.06).

**Conclusion:** This study revealed that the overall prevalence of diabetic peripheral neuropathy is relatively high in Africa. Hence, diabetic peripheral neuropathy needs situation based intervention and preventive strategy depending on their country context. Furthermore, further meta-analysis study is needed to identify associated factors for the occurrence of diabetic peripheral neuropathy.

## Introduction

Diabetes mellitus is a global health care problem and financially costly. According to IDF(international diabetic federation) latest estimated data, globally about 425 million adults in 2017 were living with diabetes; by 2045 this will rise to 629 million [1]. Worldwide, diabetes directly caused 1.6 million deaths in 2015 [2]. Over the past decade, diabetes prevalence has risen faster in low- and middle-income countries than in high-income countries [3]. In Africa, by 2017, 39 million were living with diabetes;by 2045 this will increase to 82 million [1]. Diabetes caused at least 727 billion dollars in health expenditure in 2017, about 12% of total spending on adults [1].

Morbidity and mortality in patients with diabetes mellitus (DM) is attributed to the micro-vascular and macro-vascular complications [4]. Diabetic Peripheral neuropathy (DPN) is among micro-vascular complications of diabetes that make patients prone to ulceration and amputation. DPN is common and frequent cause of morbidity and disability [5]. The Toronto consensus meeting defined DPN as asymmetrical, length-dependent sensorimotor polyneuropathy attributed to metabolic, micro vessel alterations as a result of a background long-standing hyperglycaemia and metabolic derangements[6]. Likewise, diabetic neuropathy leads to change in diabetic nerve is delayed nerve conduction velocity and the earliest histological change is segmental demyelination [7].

The prevalence of diabetic peripheral neuropathy is varying widely in the literature. This is due to differences in the diagnostic criteria employed, types of diabetes, the different methods of patient selection and the sample size [8, 9]. However, it has been estimated to be 8.4% in china [10], 48.1% in Sri Lankan[11], 29.2% in India [12], 56.2% in Yemen [9], 39.5% Jordan [13],71.1% in Nigeria [14], 16.6% in Ghana [15],and 29.5% in Ethiopia [16].

Owing to the fact that peripheral nerve damage in diabetic patients is mostly irreversible, prevention of its occurrence has been the focus, and this has led to continued search for modifiable risk factors [17]. Current studies suggest that risk factors for diabetic peripheral neuropathy include age, gender, duration of diabetes, the presence micro-vascular complications, hypertension, residence, body mass index, HbA1c, alcohol intake, hyperglycaemia, cigarette smoking, level of physical activity and marital status [12, 18-25].

Patients with peripheral neuropathy might suffer from loss or absence of protective sensation in the lower extremities leading to balance problem[26], risk of foot ulceration[22], pain and frequently disrupts sleep [27], cardiovascular morbidity and mortality [19], reduced quality of life [28], and increase costs of treatment[29]. According to reports in the literature, appropriate interventions and screening can reduce ulcers by 60% and amputations by 85% in those with high-risk diabetic neuropathy[30]. Although most previous studies have been conducted to assess the magnitude of diabetic retinopathy, diabetic foot ulcer, and diabetic nephropathy the magnitude of diabetic peripheral neuropathy among patient with diabetes mellitus remains unknown. Hence, the objective of this systematic review and meta-analysis was to estimate the pooled prevalence of diabetic peripheral neuropathy among patients with diabetes mellitus in Africa. Finding from the current study would serve as benchmark for policy-makers to implement appropriate preventive measure and to alleviate the pressing problem of diabetic peripheral neuropathy.

## Methods

### Search strategy and database

To extract all relevant literature, electronic databases such as PubMed, Google Scholar, Africa journal of online, Scopus, Web-science, WHO afro library and Cochrane and other databases were searched. In addition, a hand search of gray literature and other related articles were deployed to identify additional relevant research. This search employed articles published from 1^st^ January/ 2000 to 22^th^ August, / 2019. The search was conducted using the following MeSH and free-text terms: “peripheral neuropathy”, “diabetic neuropathy”, “diabetic polyneuropathy”, “diabetes mellitus”, and “Africa”. Boolean operators like “AND” and “OR” were used to combine search terms.

### Eligibility criteria

Studies were included if they met the following criteria: (1) All observational studies, which reported the prevalence of diabetic peripheral neuropathy;(2) articles published in peer reviewed journals and gray literature:(3) published in the English language between 2000 to 2019;and (4) studies conducted in Africa. Studies were excluded on any one of the following conditions: (1) studies which were not fully accessed; (2) studies with duplicated citation;(3) studies with poor quality score as per stated criteria;(4) articles in which fail to determine the outcomes (diabetic peripheral neuropathy);(5) peripheral neuropathy other than diabetes mellitus; and (6) Diabetic patients presenting with HIV or tuberculosis on treatment or chemotherapy.

### Selection and quality assessment

Data were extracted using standardized data extraction format prepared in a Microsoft excel by three independent authors. The extracted information from the literature included the name of author’s, year of publication, study area, study design, sample size, data collection year, data collection method, reported prevalence and its 95% confidence interval. The quality of each included study was assessed using Newcastle-Ottawa scale [31]. Studies were included in the analysis if they scored ≥5 out of 10 points in three domains of the equally weighted ten modified NOS components for cross-sectional study. Likewise, the quality score of each study was extracted from each incorporated article by three independent authors. Any disagreements at the time of data abstraction were resolved by discussion and consensus.

Additional file 1: Table S1. Methodological quality assessment of cross-sectional studies using modified Newcastle - Ottawa Scale (NOS).

### Statistical analysis

To obtain the pooled effect size, a meta-analysis using weighted inverse variance random-effects model was performed. Heterogeneity across the included studies was checked using the chi-square based Q test and the I^2^ statistics test [32]. The result of Q test was regarded to be statistically significant at p-value < 0.1. Meta-regression and sub group analyses were performed to investigate the sources of heterogeneity. Publication bias was assessed by visual inspection of a funnel plot. In addition, Egger test was conducted and a p<u>□<□0</u>.05was considered statistically significant for the presence of publication bias [33, 34]. Sensitivity analysis was employed to see the effect of single study on the overall estimation. The meta-analysis was performed using the STATA version 14 statistical software for Windows. The result was presented in the form of table and figures.

### Data synthesis and reporting

We analysed the data to estimate the pooled prevalence of diabetic peripheral neuropathy. Results were presented using forest plots. The result of this review were reported based on the Preferred Reporting Items for Systematic Reviews and Meta-Analyses (PRISMA) guideline [35]. (Supplementary file 2-PRISMA checklist) and, it is not registered in the Prospero database.

## Result

### Search results

Totally, 1,278 studies were retrieved, of which, 1,261 studies were found from six international databases and the remaining 17 were through manual search. Databases includes; PubMed (161), Scopus (53), Google scholar (507), WHO afro library (3), Cochrane reviews (7), and Africa online journal (530). Out of them, 659 duplicate records were identified and removed. From the remaining 619 articles, 492 articles were excluded after reading of titles and abstracts based on the pre-defined eligible criteria. Finally, 127 full text articles were assessed for eligibility criteria. Based on the pre-defined criteria and quality assessment, only 23 articles were included for the final analysis (Figure 1

**Figure 1.**
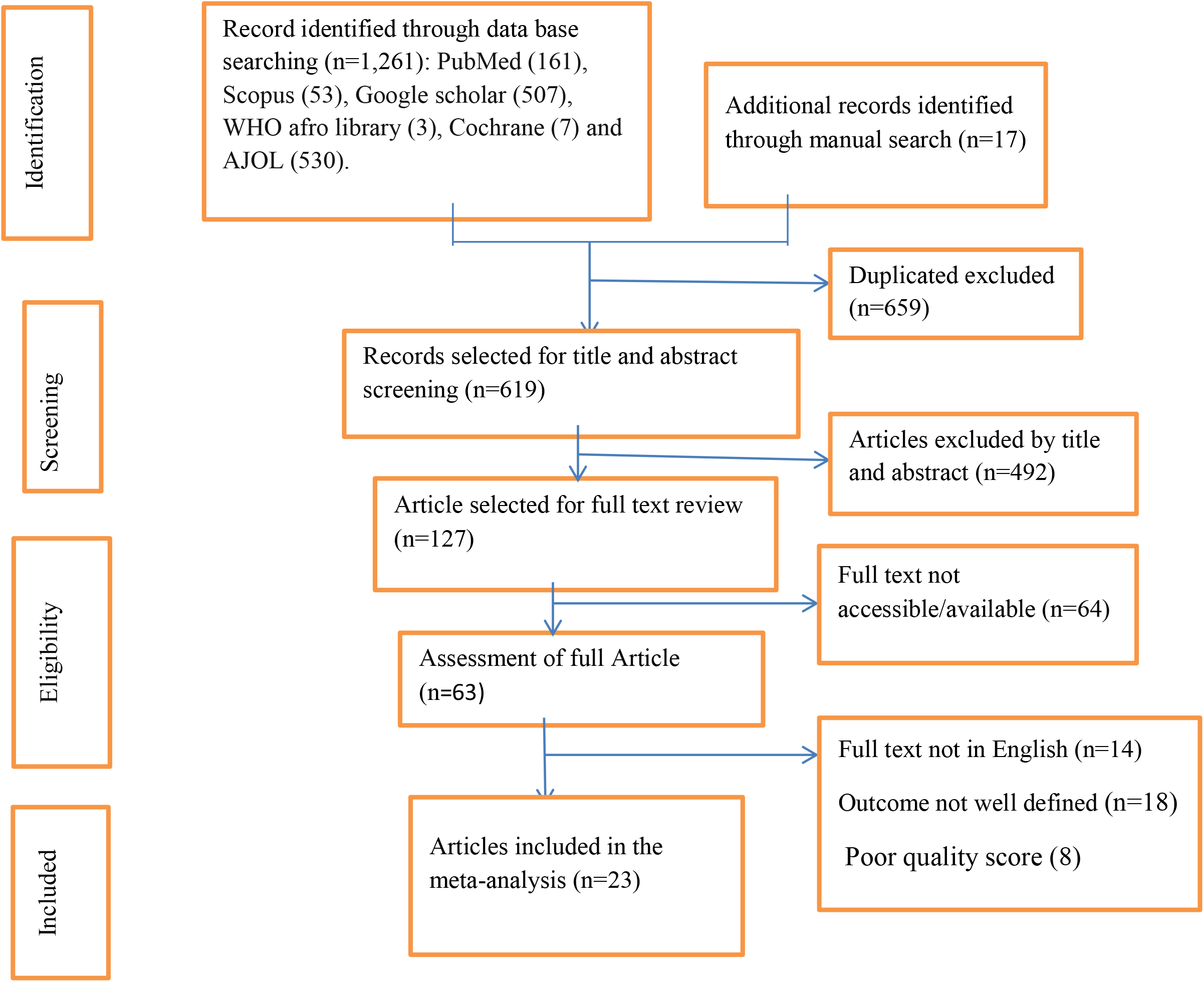
PRISMA flow chart of study selection

### Baseline characteristic of the included studies

A total of 23 studies with 269,691 participants were included in this meta-analysis. Overall information regarding the prevalence was obtained from various areas across Africa: 10 studies from Nigeria [14, 18-20, 22, 36-40], 4 article from Ethiopia[16, 24, 41, 42], 2 studies from Cameroon[23, 43], 2 article from Sudan[44, 45], 2 research paper from Egypt[46, 47],the remaining was from Ghana [15],Uganda [48],and Tanzania [49]. The highest prevalence of diabetic peripheral neuropathy (83.4%) and the lowest (7.5%) were reported from Nigeria. The studies varied substantially in size, the one having 50 patients, while the other enrolled 524 patients. Moreover, based on modified Newcastle Ottawa quality score assessment all 23 article fulfil the required quality score (Table 1).

**Table 1.**
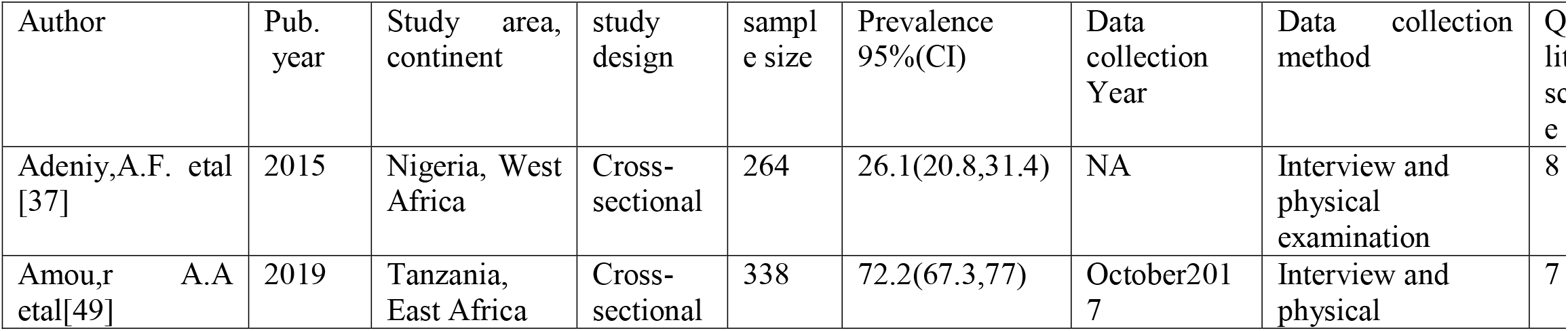
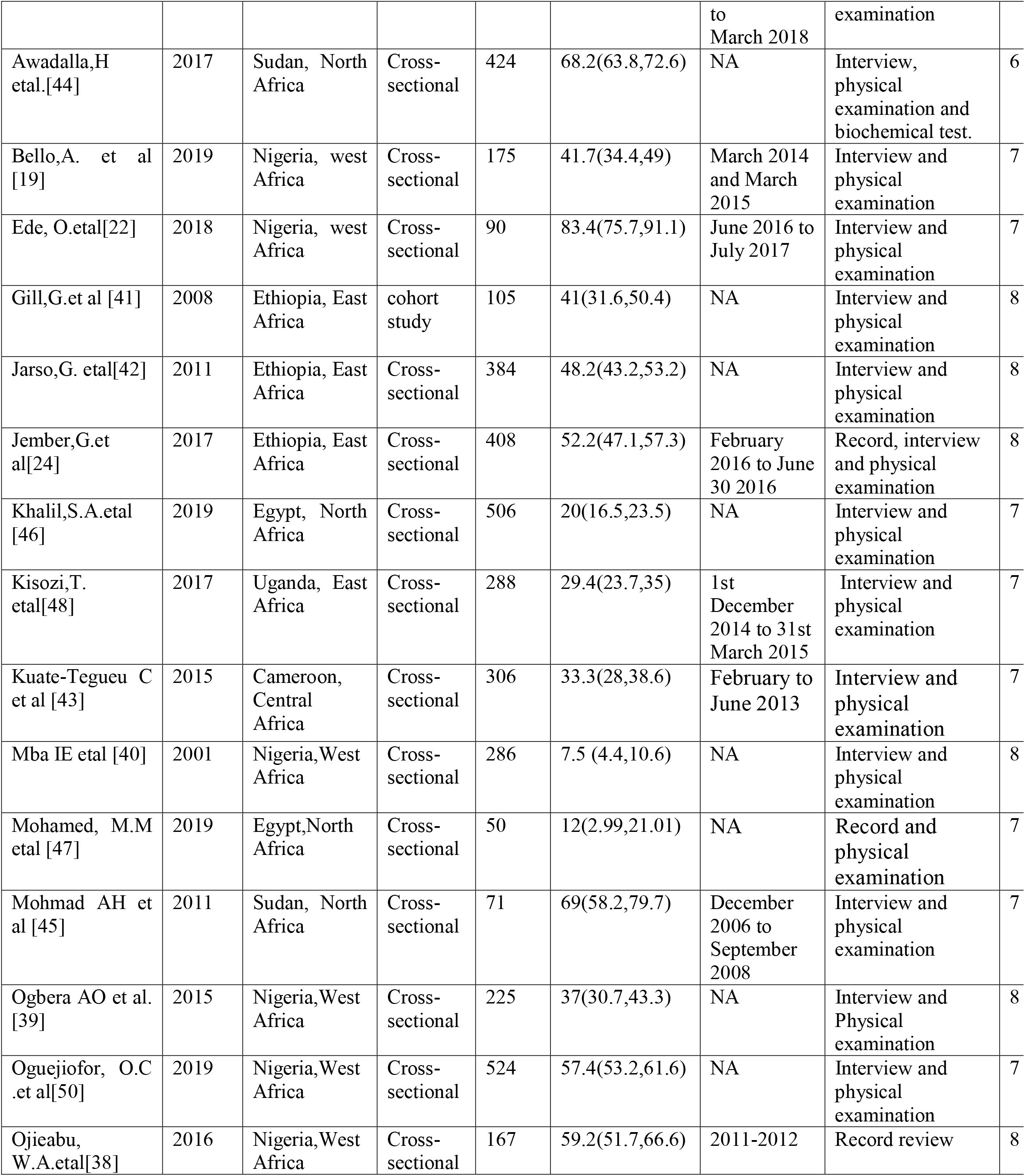
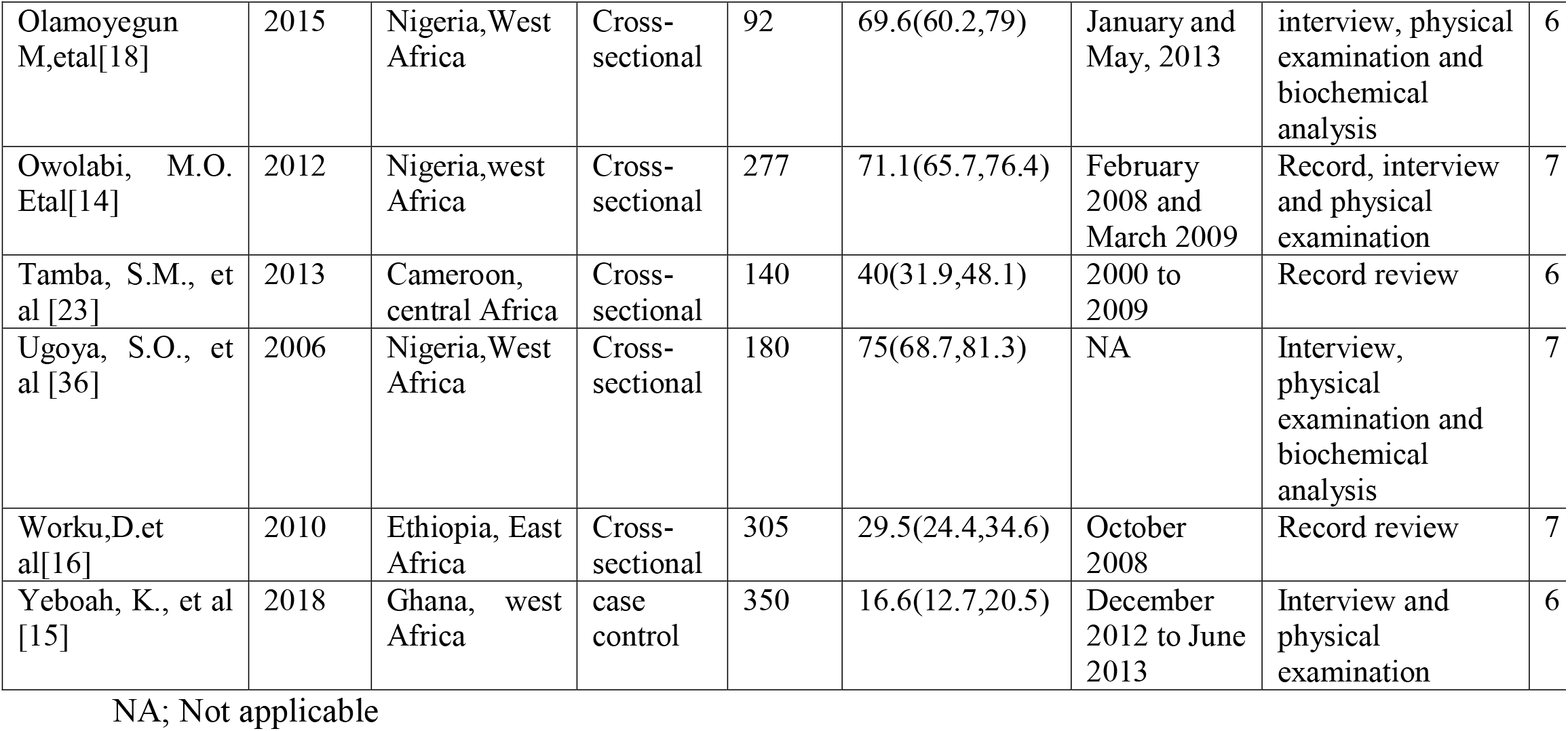
Characteristics of studies included in the meta-analysis of diabetic peripheral neuropathy in Africa.

### Prevalence of diabetic peripheral neuropathy

The result of this meta-analysis using random effects model showed that the pooled prevalence of diabetic peripheral neuropathy was 46% (95% CI: 36.21-55.78) (Figure 2) with high significant level of heterogeneity was observed (I^2^ = 98.7%; p≤0.001).

**Figure 2.**
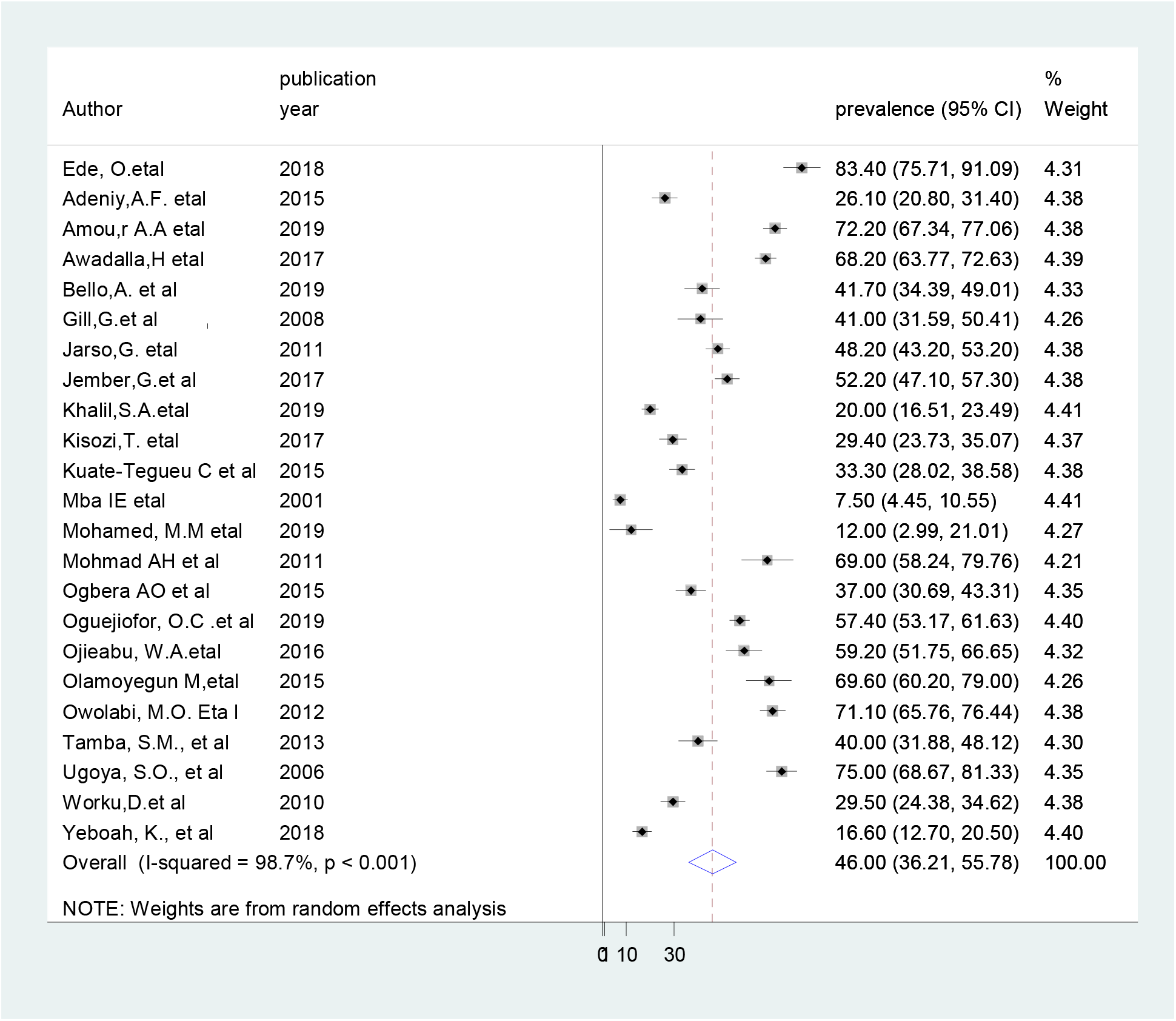
Forest plot showing the pooled prevalence of diabetic peripheral neuropathy

### Sub group analysis

In order to validate the presence of significant heterogeneity within and between the primary studies require the need to conduct subgroup analysis. As a result, in order to ascertaining the sources of heterogeneity we had deployed sub group analysis by using study area. The finding of subgroup analysis using study area showed that the highest magnitude of diabetic peripheral neuropathy was observed a study conduct in West Africa 49.4% (95% CI: 32.74, 66.06) (Figure 3).

**Figure 3.**
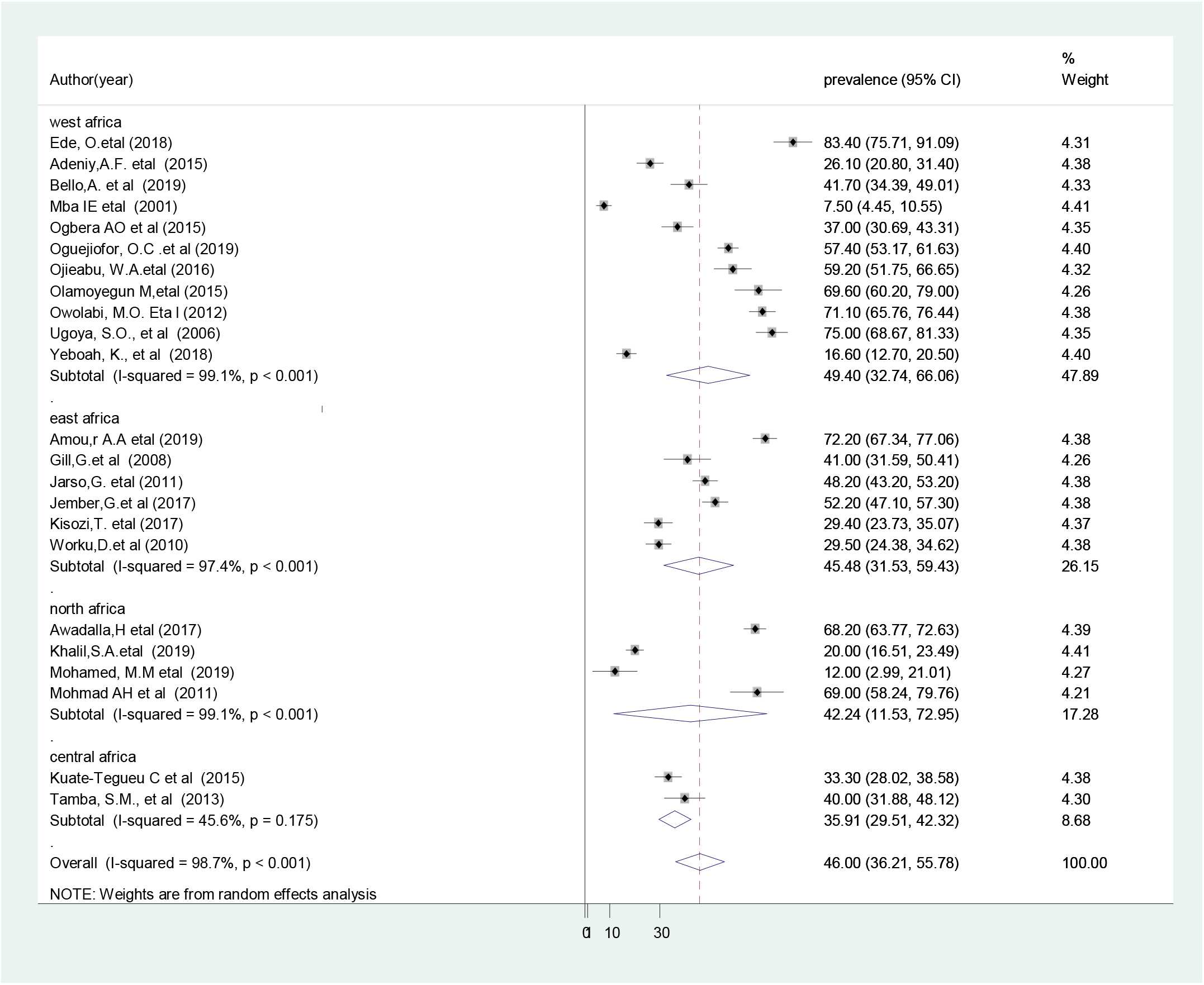
Forest plot of sub group analysis based on the country where the studies conducted.

### Meta- regression analysis

To investigate the possible source of variation across the included studies, we have performed meta-regression by using publication year, and sample size as covariate of interest. However, the result of the meta-regression analysis showed that both covariates were not statistically significant for the presence of heterogeneity (Table 2).

**Table 2.** Meta-regression analysis for the included studies to identify source of heterogeneity

| Covariate (source) | Coef. | Std. err. | t –value | P-value | 95% Conf. Interval |
| --- | --- | --- | --- | --- | --- |
| Publication year | 0.014 | 0.295 | 0.05 | 0.962 | -0.607, 0.635 |
| Sample size | -0.001 | 0.010 | -0.10 | 0.924 | -0.022, 0.020 |

### Publication bias

To identify the presence of publication bias funnel plot, and egger’s was performed. The visual inspection of the funnel plots showed asymmetrical distribution, which indicated the evidence for publication bias (Figure 4). Likewise, asymmetry of the funnel plot was statistically significant as evidenced by egger test (P = 0.024).

**Figure 4.**
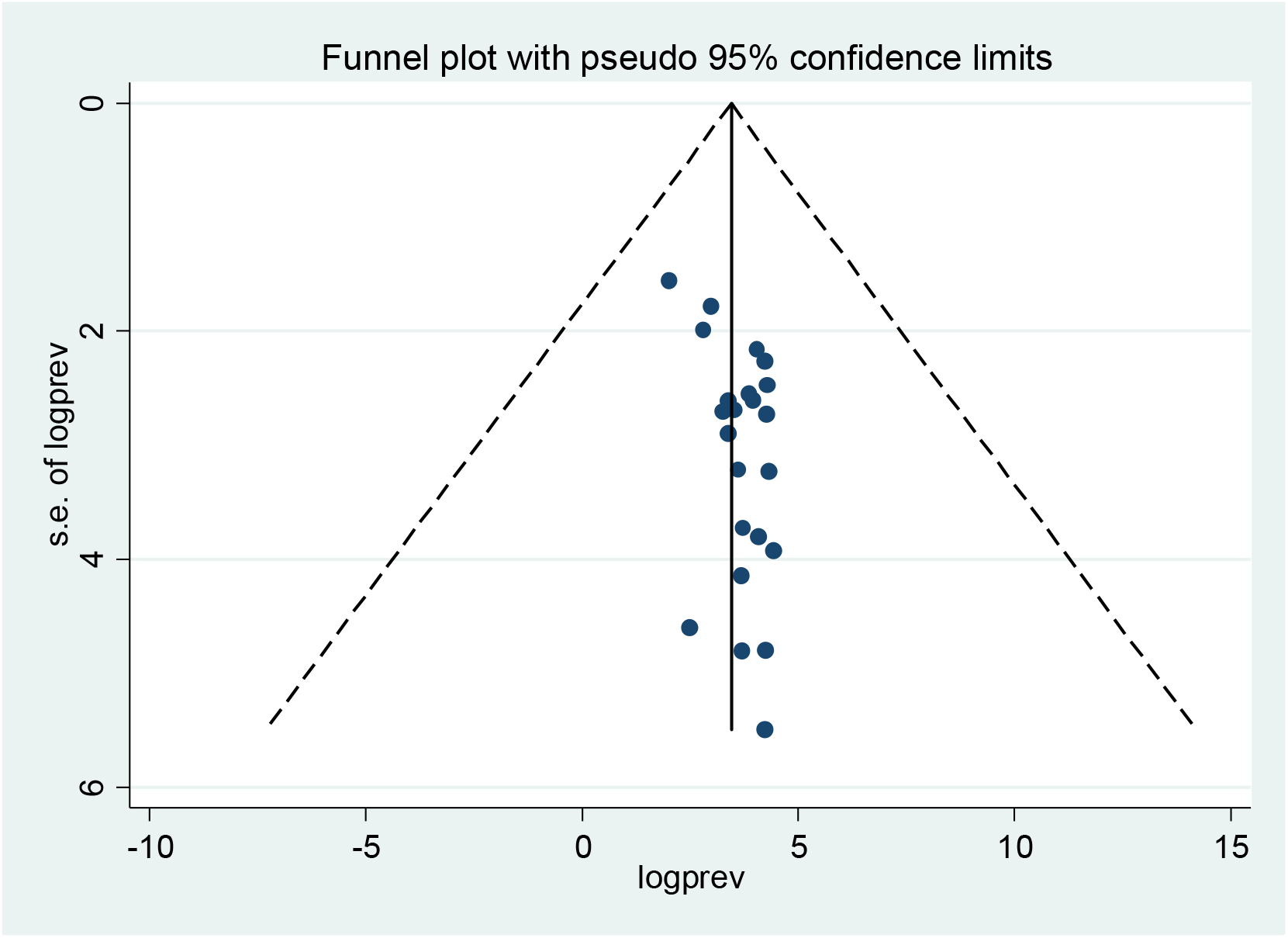
Funnel plot the presence of publication bias among 23 included studies

### Sensitivity analysis

We have also conducted sensitivity analysis, to evaluate the effect of individual study on the pooled effect size. The finding of sensitivity analyses using random effects model revealed that no single study affect the overall magnitude of diabetic peripheral neuropathy (figure 5).

**Figure 5.**
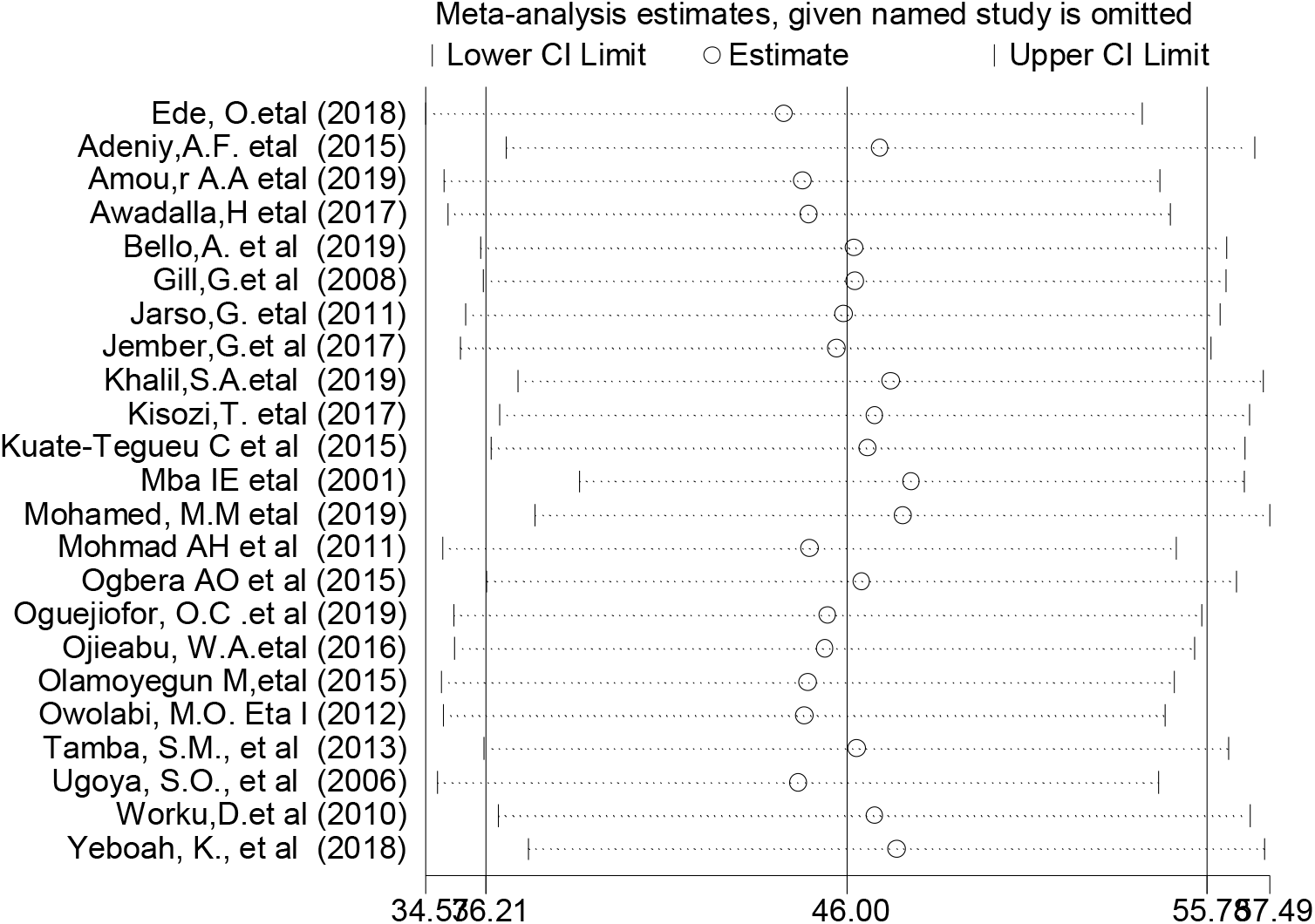
Result of sensitivity analysis of the 23 studies

## Discussion

In this systematic review and meta-analysis, the overall prevalence of diabetic peripheral nuropathy was 46%. This finding was comparable with the finding from a systematic review and meta-analysis conducted in Iran, which reported that 53% of the patients with diabetes mellitus developed diabetic peripheral neuropathy [51].On the other hand, the finding was higher than a report from a systematic review and meta-analysis in developed countries, which showed that the prevalence of diabetic peripheral neuropathy was 35.78% [52]. This variation could be different diagnostic criteria of diabetic neuropathy, early diagnosis and treatment in the developed countries.

The studies, which included in this review, had very varied in the prevalence of diabetic peripheral neuropathy that they reported from 7.5% [40] to 83.4% [22]. The result of the subgroup analysis based on study area showed that the highest pooled prevalence of diabetic peripheral neuropathy was observed from studies done in West Africa 49.4% (95% CI: 32.74, 66.06) and the lowest was observed in Central Africa 35.9% (95% CI: 29.51, 42.32). This diversity could be different diagnostic criteria of diabetic neuropathy, health care service delivery, duration and severity of diabetes. The findings of this meta-analysis have implications for clinical practice. Estimating the pooled prevalence of diabetic peripheral neuropathy provides updated evidence to develop prevention strategy, serves as key indicators of patient safety, reflect the quality of healthcare service, and appropriate management strategy for diabetic peripheral neuropathy patients.

This systematic review and meta-analysis was conducted based on PRISMA guideline for literature reviews. In addition, publication bias was quantified using Egger’s regression statistical test, and NOS was used to assess the quality of included studies. It has been the first study in Africa; the evidence could be helpful for future researchers, public health practitioners and healthcare policy-makers.

Although this meta-analysis conducted with the use of comprehensive search strategy to incorporate the studies conducted in Africa and all the included studies were observational study. In addition, the inclusion of all previously published studies is a further strength of this meta-analysis; there are some limitations that need to be considered in the future research. First, only English articles were considered; Second, this study do not identify the predictors of diabetic peripheral neuropathy; and Third, all included studies were reported hospital-based data.

## Conclusion and recommendations

This study revealed that the overall prevalence of diabetic peripheral neuropathy was relatively high in Africa. Therefore, policymakers and other concerned bodies need to give special attention to improve health care delivery for patient with diabetes mellitus and reduce the risk of peripheral neuropathy. Situation based interventions and country context specific effective preventive strategies should be developed to reduce the burden of peripheral neuropathy among patients with diabetes mellitus and to improve the overall quality of healthcare service at large. Likewise, intensive and multifactorial approach is required to combat the diabetic related complication, which focus on treatment adherence, control of comorbidity, early diagnosis and treatment. Furthermore, further research is needed to identify associated factors for the development of peripheral neuropathy among patients with diabetes mellitus.

## Data Availability

The data analyzed during the current meta-analysis is available from the corresponding author on reasonable request.

## Abbreviations

CI: Confidence Interval
DPN: Diabetic Peripheral Neuropathy
IDF: international diabetic federation
PRISMA: Preferred Reporting Items for Systematic Reviews and Meta-Analyses

## Declaration

### Ethics approval and consent to participate

Not applicable.

### Consent for publication

Not applicable.

### Authors’ contributions

WS and TY developed the protocol and involved in the design, selection of study, data extraction, and statistical analysis and developing the initial drafts of the manuscript. YA, YW and TY involved in data extraction, quality assessment, statistical analysis and revising. WS and YA prepared the final draft of the manuscript. All authors read and approved the final draft of the manuscript.

## Acknowledgements

We would like to thank all authors of studies included in this meta-analysis.

## Reference

1. IDF DIABETES ATLAS 8th edition 2017.

2. Organization WH: Guidelines on second-and third-line medicines and type of insulin for the control of blood glucose levels in non-pregnant adults with diabetes mellitus: World Health Organization; 2018.

3. Roglic G: WHO Global report on diabetes: A summary. International Journal of Noncommunicable Diseases 2016, 1(1):3.

4. Kumar KH, Kota S, Basile A, Modi K: Profile of microvascular disease in type 2 diabetes in a tertiary health care hospital in India. Annals of medical and health sciences research 2012, 2(2):103–108.

5. Genuth S, Eastman R, Kahn R, Klein R, Lachin J, Lebovitz H, Nathan D, Vinicor F: Implications of the United kingdom prospective diabetes study. Diabetes care 2003, 26:S28.

6. Tesfaye S, Boulton AJ, Dyck PJ, Freeman R, Horowitz M, Kempler P, Lauria G, Malik R, Spallone V, Vinik A: Diabetic neuropathies: Update on definitions, diagnostic criteria, estimation of severity, and treatments (Diabetes Care (2010) 33,(2285-2293)). Diabetes Care 2010, 33(12):2725.

7. Association AD: Standards of medical care in diabetes. Diabetes care 2005, 28(1):S4.

8. Edwards JL, Vincent AM, Cheng HT, Feldman EL: Diabetic neuropathy: mechanisms to management. Pharmacology & therapeutics 2008, 120(1):1–34.

9. Al Washali A, Azuhairi A, Hejar A, Amani Y: Prevalence and associated risk factors of diabetic peripheral neuropathy among diabetic patients in national center of diabetes in Yemen. International Journal of Public Health and Clinical Sciences 2014, 1(1):141–150.

10. Lu B, Hu J, Wen J, Zhang Z, Zhou L, Li Y, Hu R: Determination of peripheral neuropathy prevalence and associated factors in Chinese subjects with diabetes and pre-diabetes–ShangHai Diabetic neuRopathy Epidemiology and Molecular Genetics Study (SH-DREAMS). PLoS One 2013, 8(4):e61053.

11. Katulanda P, Ranasinghe P, Jayawardena R, Constantine GR, Sheriff MR, Matthews DR: The prevalence, patterns and predictors of diabetic peripheral neuropathy in a developing country. Diabetology & metabolic syndrome 2012, 4(1):21.

12. Bansal D, Gudala K, Muthyala H, Esam HP, Nayakallu R, Bhansali A: Prevalence and risk factors of development of peripheral diabetic neuropathy in type 2 diabetes mellitus in a tertiary care setting. Journal of diabetes investigation 2014, 5(6):714–721.

13. Khawaja N, Abu-Shennar J, Saleh M, Dahbour SS, Khader YS, Ajlouni KM: The prevalence and risk factors of peripheral neuropathy among patients with type 2 diabetes mellitus; the case of Jordan. Diabetology & metabolic syndrome 2018, 10(1):8.

14. Owolabi MO, Ipadeola A: Total vascular risk as a strong correlate of severity of diabetic peripheral neuropathy in Nigerian Africans. Ethnicity & disease 2012, 22(1):106–112.

15. Yeboah K, Agyekum JA, Owusu Mensah RN, Affrim PK, Adu-Gyamfi L, Doughan RO, Adjei AB: Arterial Stiffness Is Associated with Peripheral Sensory Neuropathy in Diabetes Patients in Ghana. Journal of diabetes research 2018, 2018.

16. Worku D, Hamza L, Woldemichael K: Patterns of diabetic complications at jimma university specialized hospital, southwest ethiopia. Ethiopian journal of health sciences 2010, 20(1).

17. Won JC, Park TS: Recent advances in diagnostic strategies for diabetic peripheral neuropathy. Endocrinology and metabolism 2016, 31(2):230–238.

18. Olamoyegun M, Ibraheem W, Iwuala S, Audu M, Kolawole B: Burden and pattern of micro vascular complications in type 2 diabetes in a tertiary health institution in Nigeria. African health sciences 2015, 15(4):1136–1141.

19. Bello A, Biliaminu S, Wahab K, Sanya E: Distal symmetrical polyneuropathy and cardiovascular autonomic neuropathy among diabetic patients in Ilorin: Prevalence and predictors. Nigerian Postgraduate Medical Journal 2019, 26(2):123.

20. Oguejiofor O, Odenigbo C, Oguejiofor C: Evaluation of the effect of duration of diabetes mellitus on peripheral neuropathy using the United Kingdom screening test scoring system, bio-thesiometry and aesthesiometry. Nigerian journal of clinical practice 2010, 13(3).

21. Azura M, Adibah H, Juwita S: Risk factor of peripheral neuropathy among newly diagnosed type 2 diabetic patients in primary care clinic. International Journal of Collaborative Research on Internal Medicine & Public Health 2012, 4(11):1858.

22. Ede O, Eyichukwu GO, Madu KA, Ogbonnaya IS, Okoro KA, Basil-Nwachuku C, Nwokocha KA: Evaluation of Peripheral Neuropathy in Diabetic Adults with and without Foot Ulcers in an African Population. Journal of Biosciences and Medicines 2018, 6(12):71–78.

23. Tamba SM, Ewane ME, Bonny A, Muisi CN, Nana E, Ellong A, Mvogo CE, Mandengue SH: Micro and macrovascular complications of diabetes mellitus in Cameroon: risk factors and effect of diabetic check-up-a monocentric observational study. Pan African Medical Journal 2013, 15(1).

24. Jember G, Melsew YA, Fisseha B, Sany K, Gelaw AY, Janakiraman B: Peripheral Sensory Neuropathy and associated factors among adult diabetes mellitus patients in Bahr Dar, Ethiopia. Journal of Diabetes & Metabolic Disorders 2017, 16(1):16.

25. Liu X, Xu Y, An M, Zeng Q: The risk factors for diabetic peripheral neuropathy: A meta-analysis. PloS one 2019, 14(2):e0212574.

26. Herrera-Rangel A, Aranda-Moreno C, Mantilla-Ochoa T, Zainos-Saucedo L, Jáuregui-Renaud K: The influence of peripheral neuropathy, gender, and obesity on the postural stability of patients with type 2 diabetes mellitus. Journal of diabetes research 2014, 2014.

27. Hoffman DL, Sadosky A, Alvir J: Cross-national burden of painful diabetic peripheral neuropathy in Asia, Latin America, and the Middle East. Pain Practice 2009, 9(1):35–42.

28. Coffey JT, Brandle M, Zhou H, Marriott D, Burke R, Tabaei BP, Engelgau MM, Kaplan RM, Herman WH: Valuing health-related quality of life in diabetes. Diabetes care 2002, 25(12):2238–2243.

29. Mehra M, Merchant S, Gupta S, Potluri RC: Diabetic peripheral neuropathy: resource utilization and burden of illness. Journal of medical economics 2014, 17(9):637–645.

30. G. N: Progress in the diagnosis of diabetic peripheral neuropathy. Chinese Journal of Practical Internal Medicine 2007, 7:487–489.

31. Shea BJ, Reeves BC, Wells G, Thuku M, Hamel C, Moran J, Moher D, Tugwell P, Welch V, Kristjansson E: AMSTAR 2: a critical appraisal tool for systematic reviews that include randomised or non-randomised studies of healthcare interventions, or both. Bmj 2017, 358:j4008.

32. Borenstein M, Hedges LV, Higgins JP, Rothstein HR: Introduction to meta-analysis: John Wiley & Sons; 2011.

33. Egger M, Davey-Smith G, Altman D: Systematic reviews in health care: meta-analysis in context: John Wiley & Sons; 2008.

34. Rücker G, Schwarzer G, Carpenter J: Arcsine test for publication bias in meta-analyses with binary outcomes. Statistics in medicine 2008, 27(5):746–763.

35. Liberati A, Altman DG, Tetzlaff J, Mulrow C, Gøtzsche PC, Ioannidis JP, Clarke M, Devereaux PJ, Kleijnen J, Moher D: The PRISMA statement for reporting systematic reviews and meta-analyses of studies that evaluate health care interventions: explanation and elaboration. PLoS medicine 2009, 6(7):e1000100.

36. Ugoya SO, Echejoh GO, Ugoya TA, Agaba EI, Puepet FH, Ogunniyi A: Clinically diagnosed diabetic neuropathy: frequency, types and severity. Journal of the National Medical Association 2006, 98(11):1763.

37. Adeniyi AF, Aiyegbusi OS, Ogwumike OO, Adejumo PO, Fasanmade AA: Habitual physical activity, peripheral neuropathy, foot deformities and lower limb function: characterising prevalence and interlinks in patients with type 2 diabetes mellitus. Journal of Endocrinology, Metabolism and Diabetes of South Africa 2015, 20(2):101–107.

38. Ojieabu WA, Odusan O, Ojieabu NI, Oku LM: Evaluation of prevalence of micro-and macrovascular complications among elderly Type 2 diabetes patients in a health facility. African Journal of Biomedical Research 2017, 20(2):131–135.

39. Ogbera AO, Adeleye O, Solagberu B, Azenabor A: Screening for peripheral neuropathy and peripheral arterial disease in persons with diabetes mellitus in a Nigerian University Teaching Hospital. BMC research notes 2015, 8(1):533.

40. Mba I: Diabetes Mellitus in Abia State University Teaching Hospital Aba, A 30 Months Retrospective Study. Journal of Medical Investigation and Practice 2001, 2(1):48–51.

41. Gill G, Gebrekidan A, English P, Wile D, Tesfaye S: Diabetic complications and glycaemic control in remote North Africa. QJM: An International Journal of Medicine 2008, 101(10):793–798.

42. Jarso G, Ahmed A, Feleke Y: The prevalence, clinical features and management of periphral neuropathy among diabetic patients in Tikur Anbessa and St. Paul’s Specialized University Hospitals, Addis Ababa, Ethiopia. Ethiopian medical journal 2011, 49(4):299–311.

43. Kuate-Tegueu C, Temfack E, Ngankou S, Doumbe J, Djientcheu V, Kengne A: Prevalence and determinants of diabetic polyneuropathy in a sub-Saharan African referral hospital. Journal of the neurological sciences 2015, 355(1-2):108–112.

44. Awadalla H, Noor SK, Elmadhoun WM, Almobarak AO, Elmak NE, Abdelaziz SI, Sulaiman AA, Ahmed MH: Diabetes complications in Sudanese individuals with type 2 diabetes: overlooked problems in sub-Saharan Africa? Diabetes & Metabolic Syndrome: Clinical Research & Reviews 2017, 11:S1047–S1051.

45. Mohmad AH, Hassan A: Correlation between retinopathy, nephropathy and peripheral neuropathy among adult Sudanese diabetic patients. Sudan Journal of Medical Sciences 2011, 6(1).

46. Khalil SA, Megallaa MH, Rohoma KH, Guindy MA, Zaki A, Hassanein M, Malaty AH, Ismael HM, Kharboush IF, Kafash E: Prevalence of chronic diabetic complications in newly diagnosed versus known type 2 diabetic subjects in a sample of Alexandria population, Egypt. Current diabetes reviews 2019, 15(1):74–83.

47. Mohamed MM, Ali RA, Hamdoon MN: Frequency and Determinants of Peripheral Neuropathy in Diabetic Children in Sohag, Egypt. Journal of Behavioral and Brain Science 2019, 9(4):184–194.

48. Kisozi T, Mutebi E, Kisekka M, Lhatoo S, Sajatovic M, Kaddumukasa M, Nakwagala FN, Katabira E: Prevalence, severity and factors associated with peripheral neuropathy among newly diagnosed diabetic patients attending Mulago hospital: a cross-sectional study. African health sciences 2017, 17(2):463–473.

49. Amour AA, Chamba N, Kayandabila J, Lyaruu IA, Marieke D, Shao ER, Howlett W: Prevalence, Patterns, and Factors Associated with Peripheral Neuropathies among Diabetic Patients at Tertiary Hospital in the Kilimanjaro Region: Descriptive Cross-Sectional Study from North-Eastern Tanzania. International Journal of Endocrinology 2019, 2019.

50. Oguejiofor OC, Onwukwe CH, Ezeude CM, Okonkwo EK, Nwalozie JC, Odenigbo CU, Oguejiofor CB: Peripheral neuropathy and its clinical correlates in Type 2 diabetic subjects without neuropathic symptoms in Nnewi, South-Eastern Nigeria. Journal of Diabetology 2019, 10(1):21.

51. Sobhani S, Asayesh H, Sharifi F, Djalalinia S, Baradaran HR, Arzaghi SM, Mansourian M, Rezapoor A, Ansari H, Masoud MP: Prevalence of diabetic peripheral neuropathy in Iran: a systematic review and meta-analysis. Journal of Diabetes & Metabolic Disorders 2014, 13(1):97.

52. de Souza LR, Debiasi D, Ceretta LB, Simões PW, Tuon L: Meta-Analysis And Meta-Regression Of The Prevalence Of Diabetic Peripheral Neuropathy Among Patients With Type 2 Diabetes Mellitus. International Archives of Medicine 2016, 9.

